# Knowledge, Attitudes, and Fear of COVID-19 during the Rapid Rise Period in Bangladesh

**DOI:** 10.1101/2020.06.17.20133611

**Authors:** Mohammad Anwar Hossain, K M Amran Hossain, Lori Maria Walton, Zakir Uddin, Md. Obaidul Haque, Md. Feroz Kabir, S. M. Yasir Arafat, Mohamed Sakel, Rafey Faruqui, Ikbal Kabir Jahid, Zahid Hossain

## Abstract

**Objectives:** To determine the level of Knowledge, Attitude, and Practice (KAP) related to COVID-19 preventive health habits and perception of Fear towards COVID-19 in subjects living in Bangladesh.

**Design:** Prospective, cross-sectional survey of (n= 2157) male and female subjects, 13-90 years of age, living in Bangladesh.

**Methods:** Ethical Approval and Trial registration were obtained prior to the commencement of the study. Subjects who volunteered to participate and signed the informed consent were enrolled in the study and completed the “Fear of COVID-19 Scale” (FCS).

**Results:** Twenty-eight percent (28.69%) of subjects reported one or more COVID-19 symptoms and 21.4% of subjects reported one or more comorbidities. Knowledge scores were slightly higher in males (8.75± 1.58) than females (8.66± 1.70). Knowledge was significantly correlated with age (p<.005), an education level (p<.001), Attitude (p<.001), and urban location (p=<.001). Knowledge scores showed an inverse correlation with Fear scores (p=<.001). Eighty-three percent (83.7%) of subjects with COVID-19 symptoms reported wearing a mask in public and 75.4% of subjects reported staying away from crowded places. Subjects with one or more symptoms reported higher Fear compared to subjects without (18.73± 4.6; 18.45± 5.1).

**Conclusions:** Overall, Bangladeshis reported a high prevalence of self-isolation, positive preventive health behaviors related to COVID-19, and moderate to high fear levels. Higher Knowledge and Practice were found in males, higher education levels, older age, and urban location. “Fear” of COVID-19 was more prevalent in female and elderly subjects. Positive “Attitude” was reported for the majority of subjects, reflecting the belief that COVID-19 was controllable and containable.

**Ethical approval:** Ethical permission obtained from the Institutional review board (BPA-IPRR/IRB/29/03/2020/021) of Institute of Physiotherapy, Rehabilitation, and Research (IPRR), the academic organization of the Bangladesh Physiotherapy Association.

**WHO Trial registry:** The trial registration obtained prospectively from a primary trial registry of WHO (CTRI/2020/04/024413).

**Data Availability:** The data are available regarding this study and can be viewed upon request

## INTRODUCTION

Bangladesh is among the top 20 countries in terms of confirmed cases of COVID 19 with a positive case rate of 19.09% - 22.91% since the first of June 2020 (Mustazir & Alif, 2020). However, questions remain regarding the actual number of cases and concerns regarding the scarcity of testing facilities (The Daily Star, 2020). There are also concerns about Bangladesh’s ability to mount an effective response to the COVID-19 pandemic (The Economist, 2020 a). This newspaper also states (2020,b), Bangladesh is a developing economy, and is largely dependent on remittances, readymade garments, and small trades. The country is in the mid-phase of a few financial megaprojects. Natural calamities and COVID-19 pose challenges for the Bangladeshi government and its residents at home and abroad (Hasina & Verkooijen, 2020). Due to the economic concerns, Bangladesh did not impose a countrywide lockdown. Instead the authority sub-sectioned the country into red, yellow, and green based on the level of community contamination (“Covid19tracker. Bangladesh Computer Council (BCC)”, 2020). Additionally, the Government website of Corona briefing (2020) measures are being taken to improve the situation raising individual awareness by improving individual’s knowledge, attitude, and practices which has helped alleviate unnecessary fears and social stigmas.

Battling with the COVID-19 pandemic is a lengthy process and requires the combined effort of individuals and the government; adequate testing, isolation, and supportive treatment provision are the best ways to overcome the pandemic (Adhikari et al., 2020). There is ongoing research to find the vaccine, but measures to raise the general population’s knowledge and implementing recommended health practices are some of the best policies to combat COVID 19 (Watkins, 2020).

The World Health Organization (2020) stated that only 15% of cases were projected to have severe symptoms and one-third of the severe cases needing critical care; the main priority of the World Health Organization (WHO) is to mobilize the resources to improve community healthcare practices. There is an emphasis on improving community’s receptiveness to staying home. Moreover, WHO raised concerns regarding mental health needs (World Health Organization: Regional Office for the Western Pacific, 2020). Mental health needs, related to COVID-19, are emerging regardless of age, occupation, and education and are related to isolation, financial uncertainty, quarantine effect, excessive online operation, gaming, physical inactivity, insomnia, anxiety, depression, and fear of COVID (Ornell, Schuch, Sordi & Kessler, 2020). The study also suggests excessive fear and anxiety led individuals in China to have more physical and psychological signs even with mild to no symptoms reported.

Bangladesh reported a few cases of suicide due to extreme fear of COVID 19, with some cases showing a negative report after the real time polymerase chain reaction (RTPCR) test, postmortem (Mamun & Griffiths, 2020). Bangladesh responded relatively early on March 2020, with no cases for nearly a week. Subsequent arrival of travelers from Italy who defied quarantine regulations could be the source of the virus (Anwar, Nasrullah & Hosen, 2020). In addition, religious gatherings and the lack of travel restrictions are considered the primary reason for the sharp upward projection (Chowdhury, 2020) in COVID cases. A population-based study was required to determine general knowledge about the disease and what practices were being taken by Bangladeshi individuals to combat COVID.

Fear is thought to be one of the main contributors towards mass anxiety and depression (Ornell, Schuch, Sordi & Kessler, 2020) it has been shown to predict a poor response to overall health, insomnia, and suppression of immunity. Other influencing factors for anxiety and depression include: occupation, knowledge, attitudes, and practice of health related habits, and other environmental indicators.

The study objectives were to determine the level of knowledge, attitude, and practice related to COVID-19 preventive health habits and underlying fear towards COVID 19 in the Bangladeshi population and how they are affected by socio-demographics factors.

## METHODOLOGY

### Study Design and Participants

This study was a prospective cross-sectional survey conducted online through a structured questionnaire. Both male and female Bangladeshi subjects, ages 13 to 80 years that were able to respond to the questionnaire were eligible for the study. Subjects with a challenged cognitive ability or an inability to communicate were excluded from the survey.

### Questionnaire

A structured questionnaire was designed to fulfill the objectives of the study. Knowledge, attitude, and practice were measured using a survey questions used in a study during the rapid raise period in China (Zhong et al., 2020). To determine fear, the “Fear of COVID 19 Scale” (FCS) has been employed, which is reported to be valid and reliable to measure fear attributed to the Coronavirus disease (Ahorsu et al., 2020). The questionnaire was complied with forward and backward translation into Bangla and a pilot study was completed before commencement of the research.

### Ethical Issues and Trial registration

Ethical permission obtained, participation was voluntary, consent has been taken and confidentiality of the information is assured. The trial registration obtained prospectively from a primary trial registry of WHO.

### Data Collection procedure

From March to April 2020, the questionnaire was disseminated online and through email and social media targeting students, professionals and public groups of Facebook and appealed to fill themselves, their family members and neighbors. A video tutorial supplied in addition to ensure appropriate response; In case of family members who are illiterate, another member assisted in responding the questionnaire. The survey was requested to be sent back after completion. 3500 questionnaires were sent, and 2200 questionnaires were returned answered. Participation was voluntary, and a written consent form was supplied with the questionnaire. The data auditor found 2157 responses were eligible to be included in the study and analyzed.

### Statistical Analysis

Descriptive statistics were employed for correct answers to knowledge and diverse attitude and practices were presented. Knowledge, fear scores, attitude and practice variables of respondents were presented and compared with independent sample t-test, one-way analysis of variance (ANOVA) or Chi-square test depending on the nature of the data. Multivariate linear regression analysis, using demographic variables as predictor variables and knowledge and fear scores as outcome variables were analyzed for significant associations. Binary logistic regression analysis was used to identify factors related to attitude and practice. Data analysis completed using the Statistical Package of social sciences (SPSS) version 20.0. The alpha level of significance was set at <.05.

## RESULTS

### Socio-demographics

Among 2157 respondents, 1166 (54.1%) were male and 991 (45.9%) were female. The mean population age was 33.48±14.65 years. Participant’s ages ranged from 13 years to 88 years and the majority of the respondents were ages 21-30 years (33.1%). Respondents were categorized as Adolescent (10-20 years), Young (21-40 years), Young adult (41-60 years), and Elderly (above 60 years). There was a larger response in higher secondary education (32.3%) and graduates (27.6). 11.2% of respondents were either undergoing primary education or reported low levels of literacy. Respondents were from all divisions of Bangladesh, the highest response was from Dhaka (23%), and lowest form Sylhet (4.5%). Majority of the respondents were non-public servant (93%), 4% were healthcare professionals, 13.4% worked or did business in a crowded place, 13.4% were students and 7.7% of the respondents reported that a relative, colleagues or a neighbor had been diagnosed with COVID 19. Other socio-demographic profiles are described in Table 1.

**Table 1:** Relationship of demographic characteristics with Knowledge and fear

| Characteristics |  | Number of participants | Knowledge score | t/F | p-value<br>(2 tailed) | Fear Score | t/F | p-value<br>(2 tailed) |
| --- | --- | --- | --- | --- | --- | --- | --- | --- |
| Gender <sup>a</sup> | Male | 1166 (54.1%) | 8.75± 1.58 | 1.159 | .247 | 18.07± 4.94 | -4.647 | .000*** |
|  | Female | 991 (45.9%) | 8.66± 1.70 |  |  | 19.07± 5.04 |  |  |
| Age <sup>b</sup> | 13-20 years | 440 (20.4) | 8.59± 1.9 | 2.318 | .024 * | 18.45± 5.15 | .794 | .593 |
|  | 21-30 years | 713 (33.1) | 8.81± 1.54 |  |  | 18.26± 4.99 |  |  |
|  | 31-40 years | 355 (16.5) | 8.55± 1.74 |  |  | 18.74± 5.35 |  |  |
|  | 41-50 years | 327 (15.2) | 8.80± 1.47 |  |  | 18.60± 4.79 |  |  |
|  | 51-60 years | 226 (10.5) | 8.88± 1.29 |  |  | 18.89± 4.58 |  |  |
|  | 61-70 years | 75 (3.5) | 8.33± 1.83 |  |  | 19.00± 4.89 |  |  |
|  | 71-80 years | 18 (.8) | 8.44± 1.79 |  |  | 19.44± 4.96 |  |  |
|  | 80-90 years | 3 (.1) | 8.67± 1.52 |  |  | 20.00± 4.35 |  |  |
| Education <sup>b</sup> | Illiterate | 58 (2.7) | 8.83± 2.0 | 6.388 | .000*** | 19.81± 3.69 | 3.284 | .006** |
|  | primary education | 183 (8.5) | 8.17± 2.13 |  |  | 18.68± 5.22 |  |  |
|  | SSC | 423 (19.6) | 8.54± 1.85 |  |  | 19.15± 5.25 |  |  |
|  | HSC) | 696 (32.3) | 8.79± 1.63 |  |  | 18.51± 4.88 |  |  |
|  | Graduation | 596 (27.6) | 8.86± 1.31 |  |  | 18.14± 4.84 |  |  |
|  | Post-graduation | 201 (9.3) | 8.82± 1.21 |  |  | 17.99± 5.39 |  |  |
| Geography <sup>b</sup> | Barisal | 203 (9.2) | 8.88± 1.16 | 8.104 | .000*** | 18.75± 5.39 | 6.484 | .000*** |
|  | Chittagong | 334 (15.2) | 9.17± 1.24 |  |  | 18.79± 4.37 |  |  |
|  | Dhaka | 499 (23) | 8.51± 2.09 |  |  | 19.15± 5.02 |  |  |
|  | Mymensingh | 103 (6) | 8.90± 1.24 |  |  | 18.42± 4.50 |  |  |
|  | Khulna | 302 (14) | 8.46± 1.62 |  |  | 18.96± 4.72 |  |  |
|  | Rajshahi | 408 (18.7) | 8.56± 1.45 |  |  | 18.39± 4.75 |  |  |
|  | Rangpur | 207 (9.4) | 8.97± 1.68 |  |  | 17.10± 5.59 |  |  |
|  | Sylhet | 101 (4.5) | 8.46± 1.64 |  |  | 16.51± 6.28 |  |  |
| Public service <sup>a</sup> | Public Servant | 150 (7) | 8.93± 1.52 | 1.738 | .082 | 17.68± 4.97 | -2.16 | .031 * |
|  | Non-public servant | 2007 (93) | 8.69± 1.65 |  |  | 18.60± 5.01 |  |  |
| Healthcare profession <sup>a</sup> | Healthcare provider | 87 (4) | 8.99± 1.05 | 1.623 | .105 | 18.62± 4.36 | .168 | .867 |
|  | Non-Healthcare person | 2070 (96) | 8.70± 1.65 |  |  | 18.53± 5.03 |  |  |
| Business or work in a crowd <sup>a</sup> | Yes | 294 (13.4) | 8.80± 1.46 | .968 | .333 | 17.91± 4.85 | -2.29 | .022 * |
|  | No | 1863 (86.4) | 8.70± 1.67 |  |  | 18.63± 5.03 |  |  |
| Study <sup>a</sup> | Students | 930 (43.1) | 8.83± 1.53 | 2.905 | .004** | 18.46± 5.11 | -.607 | .544 |
|  | Non-students | 1227 (56.9) | 8.62± 1.71 |  |  | 18.59± 4.93 |  |  |
| Diagnosed COVID 19 in community <sup>a</sup> | Yes | 166 (7.7) | 8.80± 2.02 | .704 | .093 | 18.29± 5.75 | -.650 | .516 |
|  | No | 1991 (92.3) | 8.70± 1.60 |  |  | 18.55± 4.94 |  |  |
| COVID Symptoms (last 14 days) <sup>a</sup> | Yes | 619 (28.7) | 9.04± 1.20 | 5.9 | .0001*** | 18.73± 4.6 | 1.1.4 | .002** |
|  | No | 1536 (71.3) | 8.58± 1.78 |  |  | 18.45± 5.1 |  |  |
| Comorbidity present <sup>a</sup> | Yes | 463 (21.4) | 9.03± 1.26 | 4.8 | .0001*** | 18.27± 4.8 | -1.2 | .229 |
|  | No | 1692 (78.6) | 8.62± 1.72 |  |  | 18.61± 5.04 |  |  |

<sup>a</sup>Independent t test
<sup>b</sup>One way ANOVA
\* Significant with P= <.05
\*\* Significant with P= <.005
\*\*\* Significant with P= <.001

### COVID 19 Symptoms and Comorbidity

Multiple response analyses found 28.69% of the respondents (n=619) reported one or more symptom related to COVID 19 in the last 14 days, but none reported completing a COVID-19 test during the response. The most prevalent symptoms were dry cough 19.5% (n=121), cough with sputum (14.2%), sore throat (10%), fever of more than 100 F (4.7%), anosmia or taste loss (4.5%), shortness of breath (3.7%), 4 cases of diagnosed pneumonia and 3 cases hospitalized for pneumonia were also reported. Multiple response analysis also found 463 respondents (21.4%) reported one or more comorbidities, including: diabetes (7.1%), chronic obstructive pulmonary disease (COPD) (1.9%) and heart disease or hyptertension (2.8%). Nine subjects reported a chronic neurological disability, including stroke; 20 subjects reported chronic kidney disease (CKD) and 4.6% reported chronic smoking habits.

### Knowledge

Knowledge regarding COVID was similar in both males (8.75± 1.58) and females (8.66± 1.70). There was a significant relationship found between Knowledge scores and age (p=<.005), education level (p=<.001), and geographical distribution (p=<.001). No significant difference in knowledge score was found between public servants (8.93± 1.52) and others (8.69± 1.65); between healthcare professionals (8.99± 1.05) and others (8.70± 1.65); working in a crowd (8.80± 1.46) or working alone (8.70± 1.67); or in people who reported COVID-19 positive relatives, friends or colleagues (8.80± 2.02) compared to those with non-COVID-19 associate others (8.70± 1.60). Significant differences were found between subjects with symptoms of COVID-19 (9.04± 1.20) and subjects without COVID-19 symptoms (8.58± 1.78) with statistical significance (p=<.001). Also, a significant difference (p=<.001) was found in knowledge score between subjects with comorbidities (9.03± 1.26) and subjects without comorbidities (8.62± 1.72). The detailed associations are available in Table 1.

Multiple linear regression analysis, showed a significant correlation between knowledge scores and age (*r*=.361; p<.05). Linear associations were found between knowledge and education levels, with lowest knowledge scores found in primary education compared to all other education groups (*r*=.518; p<.001). Dhaka “urban dwellers” reported significantly higher knowledge of COVID-19 symptoms and precautions, compared to subjects from rural areas of Bangladesh (p<.005). Knowledge and education levels were directly associated, with Bachelor of Science (BSc) degree holders reporting higher knowledge of COVID-19 symptoms and precautions than any other education group (p=<.005). Public servants reported higher knowledge than other non-public servants groups (p=<.05), and students reported higher knowledge of COVID-19 symptoms and precautions compared with other non-student groups (p=<.05). Subjects without symptoms showed a significant inverse relationship with knowledge compared to those with symptoms (p=<.001). (Table 2).

**Table 2:** Results of Multiple linear regression on factors associated with Knowledge and fear

| Variables | Knowledge |  |  |  |  | Fear |  |  |  |  |
| --- | --- | --- | --- | --- | --- | --- | --- | --- | --- | --- |
|  | Coefficient | Standard error | t | r | P | Coefficient | Standard error | t | r | P |
| Gender Male vs Female | -.082 | .071 | -1.159 | .25 | .247 | 1.002 | .216 | 4.647 | .10 | .001*** |
| Age 13-40 years vs others | .079 | .077 | 1.025 | .02 | .305 | .293 | .249 | 1.177 | .03 | .239 |
| Age 41-60 years vs others | -.168 | .081 | -2.074 | .04 | .038* | -.244 | .257 | -.987 | .02 | .324 |
| Age 61-90 years vs others | .361 | .171 | 2.108 | .04 | .035* | -.395 | .544 | -.713 | .02 | .476 |
| Education Primary vs others | .518 | .120 | 4.311 | .08 | .000*** | -.467 | .776 | -1.363 | .03 | .173 |
| Education secondary and higher secondary vs others | .151 | .076 | 1.99 | .009 | .046* | .206 | .355 | .581 | .04 | .561 |
| Education Bachelor and above vs others | -.206 | .073 | -2.820 | .06 | .005** | .853 | .368 | 2.318 | .06 | .021* |
| Geography Dhaka vs others | .246 | .084 | 2.984 | .06 | .003** | -.801 | .255 | -3.138 | .06 | .002** |
| Public servant vs non-public servant | -.282 | .141 | -1.993 | .04 | .046* | .849 | .432 | 1.964 | .04 | .050 |
| Business or work in a crowd vs others | -.152 | .107 | -1.425 | .02 | .154 | .729 | .326 | 2.235 | .04 | .026* |
| Students vs others | -.253 | .074 | -3.438 | .06 | .001** | .317 | .225 | 1.406 | .01 | .160 |
| Symptom present vs absent | -.474 | .137 | -3.4 | .12 | .001** | -.474 | .137 | -3.47 | .02 | .001** |
| Comorbidity present vs absent | .019 | .151 | .125 | .10 | .900 | .019 | .151 | .125 | .03 | .900 |

\* Significant with $P = < .05$
\*\* Significant with $P = < .005$
\*\*\* Significant with $P = < .001$

### Attitude

Attitude was measured in respect to “belief” on whether Bangladesh can overcome the challenge of COVID 19 or “positive synergy” towards disease control. Females reported higher belief that COVID-19 can be controlled (p=<.05). Similarly graduates, or more qualified respondents were confident that COVID-19 can be controlled (p=<.001) and also agreed that Bangladesh was capable of overcoming the challenge (p=<.001). The majority of subjects who identified as public servants in Bangladesh also reported belief that the disease was controllable (p=<.05); however, they did not believe COVID-19 would be overcome, easily (p=<.001). The subjects with a higher knowledge score of COVID-19 and higher score in the COVID-19 Fear Scale also showed higher scores in “belief” that the virus was controllable (P=<.001) and that eradication of the virus nationwide would be achieved (P=<.001). Subjects with COVID-19 symptoms and comorbidities reported higher prevalence of “belief” that the virus was both controllable and containable (p=<.001).

### Practice

Practice was measured by report of subject’s attendance at crowded areas and report of wearing a mask. The majority of female subjects in the study followed the practice of staying home (37.5%) and (36.8%) wearing a mask to prevent spread of COVID-19 (p=<.001). Similarly, graduates and above qualified personnel reported (27.9%) staying home and avoiding crowded spaces (p=<.005). The majority of the population from Dhaka followed the health advisory by staying home (55.7%) and reported wearing a mask (65.3%) (p<.001). No significant relationship was found between knowledge score and practice, but a highly significant relationship was found between the fear score and with maintaining health advisory, and between the fear score and report of mask wearing (p<.001). The majority of subjects with COVID 19 symptoms reported wearing a mask (p<.05) but also reported going to a crowded place. The majority of subjects, with comorbidities, also reported staying at home, but did not report wearing a mask (p<.005). (Table 3)

**Table 3:** Relationship among attitude and practice with demographic variables

| Variables |  | Attitude |  |  |  | Practice |  |  |  |
| --- | --- | --- | --- | --- | --- | --- | --- | --- | --- |
|  |  | A1: Believe in Control |  | A2: Bangladesh can win |  | P1: Go to crowd |  | P2: Wear Mask |  |
|  |  | Agree | Disagree | Yes | No | Yes | No | Yes | No |
| Gender | Male | 751 (34.8) | 415 (19.2) | 487 (22.6) | 679 (31.5) | 346 (16.0) | 818 (37.9) | 1012 (46.9) | 154 (7.1) |
|  | Female | 586 (27.2) | 405 (18.8)* | 394 (18.3) | 597 (27.7) | 178 (8.3) | 808 (37.5)*** | 794 (36.8) | 197 (9.1)*** |
| Age | 13-40 years | 925 (42.9) | 583 (27.0) | 595 (27.6) | 913 (42.3) | 350 (16.2) | 1158 (53.7) | 1255 (58.2) | 253 (11.7) |
|  | 41-60 years | 357 (16.6) | 196 (9.1) | 250 (11.6) | 303 (14.0) | 153 (7.1) | 400 (18.5) | 474 (22.0) | 79 (3.7) |
|  | 61-90 years | 55 (2.5) | 41 (1.9) | 36 (1.7) | 60 (2.8) | 21 (1.0) | 75 (3.4) | 77 (3.6) | 19 (.9) |
| Education | Primary | 125 (5.8) | 116 (5.4) | 63 (2.9) | 178 (8.3) | 63 (2.9) | 178 (8.3) | 201 (9.3) | 40 (1.9) |
|  | SSC and HSC | 677 (31.4) | 442 (20.5) | 452 (21.0) | 667 (30.9) | 266 (12.3) | 853 (39.5) | 929 (43.1) | 190 (8.8) |
|  | Bachelor and above | 535 (24.8) | 262 (12.1) *** | 366 (17.0) | 431 (20.0) *** | 195 (9.0) | 602 (27.9) ** | 676 (31.3) | 121 (5.6) |
| Geography | Dhaka | 264 (12.2) | 235 (10.9) | 168 (7.8) | 331 (15.3) | 69 (3.2) | 430 (19.9) | 397 (18.4) | 102 (4.7) |
|  | Other parts | 1073 (49.7) | 585 (27.1) *** | 713 (33.1) | 945 (43.8) *** | 455 (21.1) | 1203 (55.7)*** | 1409 (65.3) | 249 (11.5)** |
| Public service | Public Servant | 108 (5.0) | 42 (1.9)** | 82 (3.8) | 68 (3.2) *** | 43 (2.0) | 107 (5.0) | 130 (6.0) | 20 (.9) |
| Health Profession | Healthcare provider | 57 (2.6) | 30 (1.4) | 40 (1.9) | 47 (2.2) | 28 (1.3) | 59 (2.7) | 79 (3.7) | 8 (.4) |
| Business | Work in a crowd | 208 (9.6) | 86 (4.0)** | 113 (5.2) | 181 (8.4) | 123 (5.7) | 171 (7.9)*** | 272 (12.6) | 22 (1) *** |
| Study | Students | 560 (26.0) | 370 (17.2) | 357 (16.6) | 573 (26.6)* | 196 (9.1) | 729 (33.8)** | 755 (35.0) | 175 (8.1)** |
| Diagnosed COVID 19 | in community | 144 (5.3) | 52 (2.4) | 45 (2.1) | 121 (5.6)*** | 55 (2.5) | 110 (5.1)* | 140 (6.5) | 26 (1.2) |
| Symptoms | Present | 442 (20) | 177 (8.2)*** | 248 (11.4) | 371 (17.1) | 160 (7.4) | 459 (21.2) | 533 (24.7) | 86 (3.9)* |
| Comorbidity | Present | 350 (16.2) | 113 (5.2)*** | 188 (8.7) | 275 (12.7) | 137(6.3) | 326 (15.1)** | 396 (18.3) | 67 (3.1) |
| Total Knowledge Score |  | 8.9± 1.3 | 8.3± 2*** | 8.9± 1.2 | 8.5± 1.8*** | 8.6± 1.5 | 8.7± 1.6 | 8.7± 1.6 | 8.5± 1.7 |
| Total Fear Score |  | 17.9± 4.9 | 19.4± 5.0*** | 18.2± 4.8 | 18.7± 5.1* | 17.7± 5.4 | 18.7± 4.8*** | 18.5± 4.7 | 18.2± 6.3*** |

\* Significant with $P = < .05$
\*\* Significant with $P = < .005$
\*\*\* Significant with $P = < .001$

### Fear

Fear scores were strongly associated with gender, education and geography (p=<.001), with females reporting a higher score (19.07± 5.04)and respondents’ aged 61-70 years, 71-80 years and 80-90 years reporting a higher score of fear of contracting COVID-19 (19.00± 4.89; 19.44± 4.96; 20.00± 4.35). Dhaka urban dwellers also reported a higher status of fear than rural dwellers (19.15± 5.02). The demographic relationship of fear scores are listed in Table 1 and Figure 1. Multiple linear regression found females have fear score differences than male (p=<.001, *r*=.045). Other regression are described in Table 2. An indirect, strong, but significant relationship (p=<.001) was found between fear scores and practice of recommended health advisory habits of subjects (See Table 3). There were significant differences (p=<.005) in fear scores between subjects with a symptom and those without a symptom (18.73± 4.6; 18.45± 5.1) (Table 1). Inverse relationships were found among persons having positive COVID-19 symptoms and fear score (p=<.005, *r*=.08).

**Figure 1:**
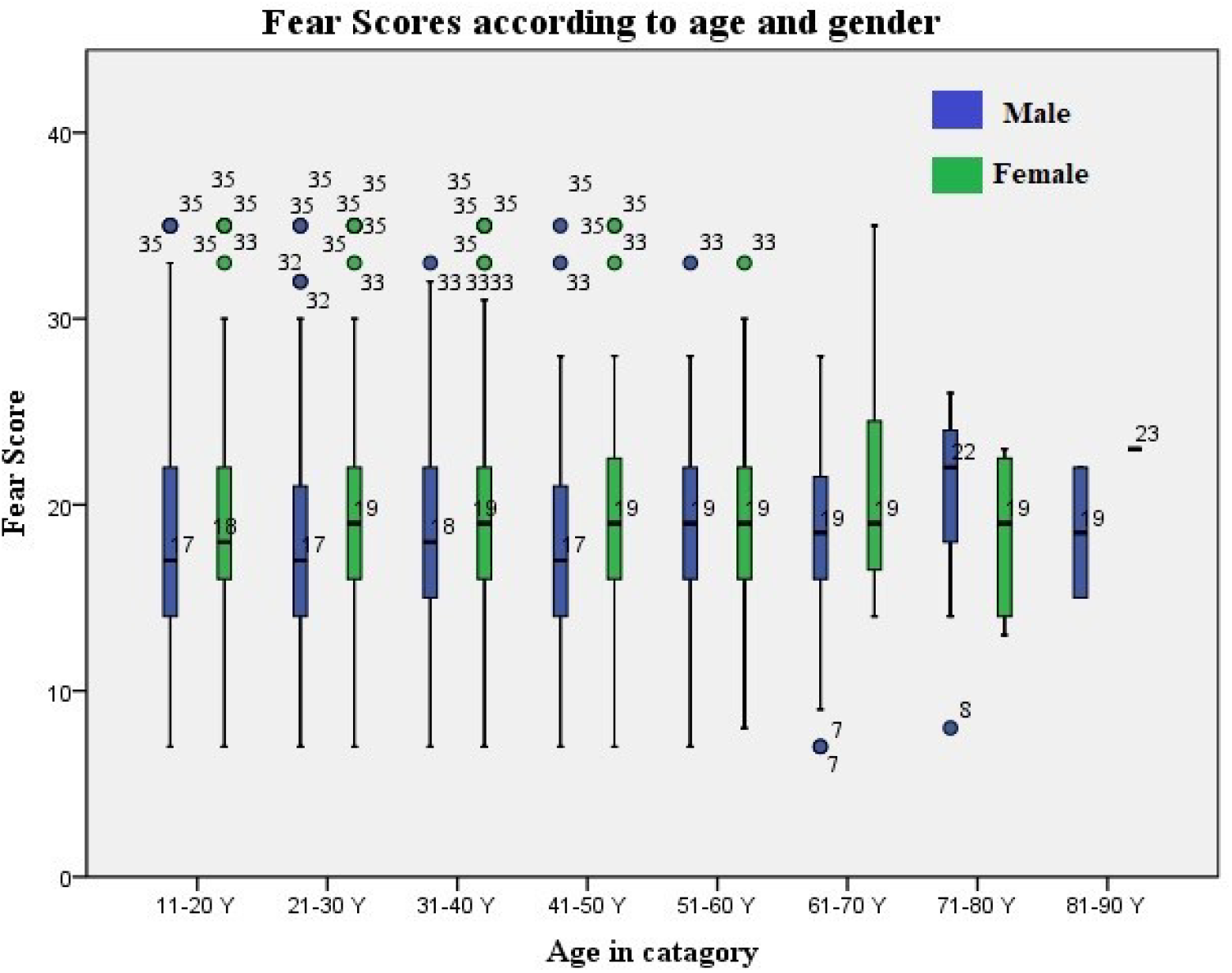
Fear Score according to age and gender

## DISCUSSION

The study intended to explore the knowledge, attitude, and practice of recommended health advice for prevention of COVID-19 and to explore the impact of fear towards contracting COVID-19 for people living in Bangladesh. There is little to no research published on this important topic for Bangladesh to date. The study covered every geographical area of Bangladesh according to administrative distribution and provided a glimpse of overview in the time frame.

Among the respondents, males (54.1%) were higher than females (45.9%); the responses per age group were 20.4% aged 13-20 years, 33.1% aged 21-30 years, 16.5% aged 31-40 years, 15.2% aged 41-50 years, 10.5% aged 51-60 years, 3.5% aged 61-70 years, .8% (n=18) aged 71-80 years and .1% (n=3) aged 80-90 years. 58 respondents (2.7%) were illiterate, 183 persons (8.5%) had only primary education, 423 respondents (19.6%) had secondary education, 696 persons (32.3%) had higher secondary degree, 596 respondents (27.6) obtained graduation degree and 201 persons (9.3%) had post-graduation degrees. The highest response of COVID-19 cases were from Dhaka (23%). The Institute for Epidemiology, Disease Control and Research (IEDCR) COVID-19 update (2020) reported that Dhaka with the majority of SARS-CoV_2 cases, and it is considered to have the most challenges regarding the level of practice of healthcare advisory precautions (Mamun, 2020). Baseline characteristics of subjects in comparative studies varied across the world. Studies in India (Roy et al., 2020), China (Zhong et al., 2020), and Egypt (Abdelhafiz et al., 2020) had more responses from females, and the USA (Clements, 2020) had more responses in males. Indian, Chinese, and Egyptian studies had similar responses of age group and education, while the USA study reported a mean age higher than our study. Our study found a satisfactory level of knowledge by gender, geography, occupation, and education (Table 1) and relatively higher fear score compared to similar studies across the world. One study, in China, showed similar scores of fear by age, with respect to knowledge, and occupation Zhong et al., 2020), while another study completed in India (Roy et al., 2020), reported 80% of people in need of mental healthcare-for COVID-19 related fear, anxiety, and depression.

Twenty-nine percent (28.69%) of the respondents (n=619) reported one or more than one symptom related to COVID 19 in the last 14 days including cough 19.5% (n=121), cough with sputum (14.2%), sore throat, (10%), fever (4.7%), anosmia or taste loss (4.5%), and shortness of breath (3.7%). The symptoms were related to COVID 19, as per CDC (“Coronavirus Disease 2019 (COVID-19) – Symptoms”, 2020). WHO South East Asia region (2020) reported the test positivity in Bangladesh to be 20%, with positive tests being reported only for a person with one or more COVID-19 related symptoms. Eleven percent (11%) of subjects reported co-morbidities, including subjects with disabilities. In addition, 4.4% of the respondents were over 60 years of age. The WHO South East Asia region (2020) country profile and Institute for Epidemiology, Disease Control and Research IEDCR COVID update (2020) states that the number of deaths are higher in elderly persons, males, and those with pre-existing co-morbidities for Bangladesh. Overall, we found a low number of elderly with symptoms, low reported levels of comorbidities, but a slightly higher rate of infection by males compared to females.

Knowledge regarding COVID-19, by subjects, was similar and satisfactory when comparing age, gender, and occupation. There were a few variations in the perspectives of occupation. Young adults, graduates, urban dwellers had more knowledge than the older adults, with lower education, living in rural areas. Several similar articles in preprint (Haque et al., 2020; Karim et al., 2020; Ferdous et al., 2020) found more than half of the respondents reported “good knowledge” of COVID-19, with age and education showing a significant linear association with knowledge. This study is similar to one study in China that found a significant relationships between knowledge and age and knowledge and educational level, with males reporting higher levels of knowledge than females regarding COVID-19 symptoms, precautions and health advisory practices (Zhong et al., 2020). However, in our study, subjects living in Bangladesh reported similar knowledge for both males and females regarding COVID-19 symptoms, precautions and advisory health practices.

Overall, a high prevalence of “positive attitudes” from the subjects towards the belief of disease control was reported. Female subjects and subjects with higher education were more likely to believe that COVID-19 can be controlled, but they doubt the ability of Bangladesh to contain it. Subjects with “good knowledge” or “high score of fear”, both were more likely to believe that COVID-19 can be controlled and that a collective effort can contain the spread of the disease. Similar studies in Bangladesh, India and China all found similar results regarding the relationship between knowledge and fear of COVID-19 (Haque et al., 2020; Ferdous et al., 2020; Roy et al., 2020; Zhong et al., 2020) regarding “practice”, our study reported similarities to previous studies across the world (Roy et al., 2020; Zhong et al., 2020; Abdelhafiz et al., 2020; Clements, 2020). Our study found (37.5%) of women reported “staying home” and 36.8% reported wearing masks in public places. The majority of the population outside of Dhaka, in the more rural regions, reported advisory staying home (55.7%) and wearing a mask (65.3%). However, no statistical relationship was found between knowledge scores and practices. This is similar to results reported in studies in both India and China. Our study did find, however, subjects with a high score of fear, and were also more likely to follow good preventive practice as recommended by the health advisory.

The fear score was significantly associated with female gender, higher education, and urban dwellers. The senior citizens aged 61-70 years, 71-80 years, and 80-90 years reported the highest score of fear compared to all categories. One study suggests that fear comes from longer duration of isolation, movement restriction, and being reactive to news and rumors from social media (Banerjee, 2020). Women, senior citizens, and young adults had limited movement, were isolated to quarantine and were attached to media. Studies report fear and stress can lead to insomnia and psychological illness (Zhong et al., 2020). However, though important as an indicator, this was not evaluated in this study, and may be considered a limitation of the study. Our study faced many challenges regarding the structured questionnaires, reporting and resources. Limitations response rates, correlation with fear and psychological issues, and completion of the questionnaire for COVID-19 positive. We recommend future studies to include information on limitation of movement, isolation and insomnia as it relates to psychological illness, as well as information regarding neurological signs and symptoms of patients and relationship to cognition and fear.

## CONCLUSION

In a resource-challenged country, like Bangladesh, individual knowledge, positive attitude, and practice of suggested precautionary and preventive health advisories are crucial to control vicious community transmission of COVID 19. The study found knowledge levels were adequate in majority of population and were directly and significantly related to higher education levels, younger age, and female gender. There were positive attitude among respondents regarding control the disease and overcome challenges of COVID-19 in Bangladesh. Majority of the population had a high score of fear and significant higher scores found in women and elderly. Surprisingly, the person having a higher score had a good practice of staying home and wearing masks. Future studies on explanatory issues related to activity, function, social issues and quality of life might add more insight studying bio-psychological impact of COVID-19 in the most densely populated country in the world.

## Data Availability

The data are available regarding this study and can be viewed upon request

## Acknowledgement

Authors acknowledges Shafin Rubayet and Ahnaf Al Mukit, research assistants for their contribution in data collection and input, also students of Bangladesh Health Professions Institute helped in collecting data.

## REFERENCES

Abdelhafiz, A., Mohammed, Z., Ibrahim, M., Ziady, H., Alorabi, M., Ayyad, M., & Sultan, E. (2020). Knowledge, Perceptions, and Attitude of Egyptians Towards the Novel Coronavirus Disease (COVID-19). Journal Of Community Health. doi: 10.1007/s10900-020-00827-7

Adhikari, S., Meng, S., Wu, Y., Mao, Y., Ye, R., & Wang, Q. et al. (2020). Epidemiology, causes, clinical manifestation and diagnosis, prevention and control of coronavirus disease (COVID-19) during the early outbreak period: a scoping review. Infectious Diseases Of Poverty, 9(1). doi: 10.1186/s40249-020-00646-x

Ahorsu, D., Lin, C., Imani, V., Saffari, M., Griffiths, M., & Pakpour, A. (2020). The Fear of COVID-19 Scale: Development and Initial Validation. International Journal Of Mental Health And Addiction. doi: 10.1007/s11469-020-00270-8

Anwar, S., Nasrullah, M., & Hosen, M. (2020). COVID-19 and Bangladesh: Challenges and How to Address Them. Frontiers In Public Health, 8. doi: 10.3389/fpubh.2020.00154

Banerjee, D. (2020). The COVID-19 outbreak: Crucial role the psychiatrists can play. Asian Journal Of Psychiatry, 50, 102014. doi: 10.1016/j.ajp.2020.102014

Chowdhury, T. (2020). Fears grow over Bangladesh’s COVID-19 response. Retrieved 14 June 2020, from https://www.aljazeera.com/news/2020/03/fears-grow-bangladeshs-covid-19-response-200323111803294.html.

Clements, J. (2020). Knowledge and Behaviors Toward COVID-19 Among US Residents During the Early Days of the Pandemic: Cross-Sectional Online Questionnaire. JMIR Public Health And Surveillance, 6(2), e19161. doi: 10.2196/19161

corona.gov.bd, a. (2020). Coronavirus Disease 2019 (COVID-19) Information Bangladesh | corona.gov.bd. Retrieved 7 June 2020, from https://corona.gov.bd/

Coronavirus Disease 2019 (COVID-19) – Symptoms. (2020). Retrieved 14 June 2020, from https://www.cdc.gov/coronavirus/2019-ncov/symptoms-testing/symptoms.html

covid-19 Update. (2020). Retrieved 14 June 2020, from https://iedcr.gov.bd/website/index.php/component/content/article/73-ncov-2019

Covid19tracker. Bangladesh Computer Council (BCC). (2020). Retrieved 7 June 2020, from http://covid19tracker.gov.bd/

Ferdous, M., Saiful Islam, M., Sikder, M., Mosaddek, A., Zegarra-Valdivia, J., & Gozal, D. (2020). Knowledge, attitude, and practice regarding COVID-19 outbreak in Bangladeshi people: An online-based cross-sectional study. doi: 10.1101/2020.05.26.20105700

Haque, T., Hossain, K., Bhuiyan, M., Ananna, S., Chowdhury, S., Ahmed, A., & Rahman, M. (2020). Knowledge, attitude and practices (KAP) towards COVID-19 and assessment of risks of infection by SARS-CoV-2 among the Bangladeshi population: An online cross sectional survey. doi: 10.21203/rs.3.rs-24562/v1

Hasina, S., & Verkooijen, P. (2020). Fighting cyclones and coronavirus: how we evacuated millions during a pandemic. The Guardian. Retrieved from https://www.theguardian.com/global-development/2020/jun/03/fighting-cyclones-and-coronavirus-how-we-evacuated-millions-during-a-pandemic

Karim, A., Akter, M., Mazid, A., Pulock, O., Aziz, T., & Hayee, S. et al. (2020). Knowledge and attitude towards COVID-19 in Bangladesh: Population-level estimation and a comparison of data obtained by phone and online survey methods. doi: 10.1101/2020.05.26.20104497

Mamun, M., & Griffiths, M. (2020). First COVID-19 suicide case in Bangladesh due to fear of COVID-19 and xenophobia: Possible suicide prevention strategies. Asian Journal Of Psychiatry, 51, 102073. doi: 10.1016/j.ajp.2020.102073

Mamun, S. (2020). Covid-19: Bangladesh records highest 45 deaths, 3,171 cases in a day. Dhaka Tribune. Retrieved from https://www.dhakatribune.com/bangladesh/2020/06/09/covid-19-record-45-deaths-and-3171-fresh-cases-in-a-day.

Mustazir, H., & Alif, A. (2020). Covid-19: Bangladesh records highest 42 deaths in a day, cases cross 65,000 mark. Dhaka Tribune. Retrieved from https://www.dhakatribune.com/health/coronavirus/2020/06/07/covid-19-bangladesh-records-highest-42-deaths-in-a-day-cases-cross-65-000-mark.

Ornell, F., Schuch, J., Sordi, A., & Kessler, F. (2020). “Pandemic fear” and COVID-19: mental health burden and strategies. Brazilian Journal Of Psychiatry, 42(3), 232–235. doi: 10.1590/1516-4446-2020-0008

Roy, D., Tripathy, S., Kar, S., Sharma, N., Verma, S., & Kaushal, V. (2020). Study of knowledge, attitude, anxiety & perceived mental healthcare need in Indian population during COVID-19 pandemic. Asian Journal Of Psychiatry, 51, 102083. doi: 10.1016/j.ajp.2020.102083.

The Daily Star. (2020). India-Pak-Bangladesh: Official Covid-19 numbers disguise undercounting. Retrieved from https://www.thedailystar.net/backpage/news/india-pak-bangladesh-official-covid-19-numbers-disguise-undercounting-1910197.

The Economist. (2020,a). Bangladesh cannot afford to close its garment factories. Retrieved from https://www.economist.com/asia/2020/04/30/bangladesh-cannot-afford-to-close-its-garment-factories

The Economist. (2020,b). Covid-19 infections are rising fast in Bangladesh, India and Pakistan. Retrieved from https://www.economist.com/asia/2020/06/06/covid-19-infections-are-rising-fast-in-bangladesh-india-and-pakistan

Watkins, J. (2020). Preventing a covid-19 pandemic. BMJ, m810. doi: 10.1136/bmj.m810

WHO South-East Asia Region. (2020). Coronavirus disease (COVID-2019) Bangladesh situation reports. Bangladesh. Retrieved from https://www.who.int/docs/default-source/searo/bangladesh/covid-19-who-bangladesh-situation-reports/who-ban-covid-19-sitrep-15-20200608.pdf?sfvrsn=c2b0efc8_4

World Health Organization. (2020). COVID-19 strategy update - 14 April 2020. Retrieved from https://www.who.int/publications/i/item/covid-19-strategy-update---14-april-2020

World Health Organization. Regional Office for the Western Pacific. (2020). Mental health and psychosocial support aspects of the COVID-19 response. Manila : WHO Regional Office for the Western Pacific. Retrieved from http://iris.wpro.who.int/handle/10665.1/14515

Zhong, B., Luo, W., Li, H., Zhang, Q., Liu, X., Li, W., & Li, Y. (2020). Knowledge, attitudes, and practices towards COVID-19 among Chinese residents during the rapid rise period of the COVID-19 outbreak: a quick online cross-sectional survey. International Journal Of Biological Sciences, 16(10), 1745–1752. doi: 10.7150/ijbs.45221

